# Association between device measured and self-reported sitting in relation to depression: the 1970 British Cohort Study

**DOI:** 10.1101/2020.03.13.20035451

**Authors:** Andrew Webster, G David Batty, Natalie Pearson, Emmanuel Stamatakis, Mark Hamer

## Abstract

**Aims:** While physical activity appears to confer protection against depression, the relationship between sedentary behaviour and mental health is uncertain. Self-reported methods provide information about context although there may be error in the quantification of sedentary behaviour. Accordingly, we examined associations of both device-measured and self-reported sedentary behaviour with depression.

**Method:** Participants (n=4,704; 52.4% Female; aged 46-48) were drawn from the 1970 British Cohort Study. Sitting time and moderate-vigorous physical activity was measured using a thigh-worn accelerometer device (ActivPAL) over a seven day period. A range of self-reported sedentary behaviours was measured to provide context. Depression diagnosis was captured using a combination of self-reported consultation with a physician and use of anti-depressant medication. Malaise inventory was used to assess depressive symptoms.

**Results:** Relative to those who spent <8 hr/d sitting, those in the highest tertile of device measured sitting (>10 hr/d) had increased odds of depression diagnosis (odds ratio= 1.48 [95% confidence interval 1.05-2.08]). There was no association between self-reported TV viewing and depression diagnosis (1.07; 0.71-1.63). We observed protective associations between moderate-vigorous physical activity and depression diagnosis (highest tertile vs. the lowest tertile; 0.70;0.49-1.00). Associations of sitting time and physical activity with depression were mutually independent. Relative to <1 hours of internet usage, 2-3 and >3 hours of internet weekday usage were associated with increased odds of depressive symptoms (1.60;1.30-1.97 and 1.63;1.32-2.03, respectively).

**Conclusion:** Device-measured sitting is associated depression diagnosis, although less consistent associations are observed with self-reported sedentary behaviours. Regular physical activity and reducing sedentary time may be beneficial for prevention of depression.

## Introduction

Common Mental Disorder (CMD) (depression and anxiety), are highly prevalent in the United Kingdom (UK) with one in six UK adults experiencing a mental health issue in any given week.[1] Furthermore it has been suggested that depression will be the world leading cause for disease burden and disability by 2030.[2] Whilst CMD has been highlighted as an important health outcome in it’s own right, CMD has been linked to an increased risk of suicide, unintentional injury and chronic disease such as cardiovascular events and selected cancers. [3–6]

There is potential to prevent depression with regular physical activity (PA) and exercise. [7] Results from a recent meta-analysis of prospective cohort studies showed regular high levels of PA was associated with lowered odds of future depressive symptoms irrespective of age and sex. [8] However, evidence for an association between sedentary behaviour (SB) and depression is less well established. SB is defined as “Any waking behaviour characterized by an energy expenditure ≤1.5 Metabolic Equivalents (METs), while in a sitting, reclining or lying posture.”[9] The existing evidence is largely based on observational data,[10,11] although evidence from a recent randomised controlled trial observed a significant increase in depressive mood when young physically active adults were placed under experimental sedentary conditions.[12]

In most studies SB has been assessed using self-reported methods. Whilst the utilisation of self-reported SB is convenient for large scale data collection and does provide important information on context, the method suffers from reporting bias.[13] Indeed, reporting bias may be partly driven by depressive symptoms as people with low mood may respond differently to questions about their lifestyle.[14] This could explain why there are some inconsistent and contrasting findings on the links between SB and depression.[15–17] Wearable devices yield more valid and reliable measurement of SB when compared to a self-report questionnaire.[18] However, accelerometery does not capture SB context that may provide important information.

To the best of our knowledge, no large scale, population-based cohort study has utilised the gold standard thigh-mounted accelerometery approach to examine associations between sitting time and depression. The aims of the current study therefore were to investigate associations of device measured sitting and self-reported sedentary behaviours with depression in a population sample of British adults.

## Method

### Sample and Study Design

The 1970 British Cohort Study (BCS70)follows the lives of people born in 1970 in a single week in England, Scotland and Wales. [19] For the present cross-sectional study we utilised data from the age 46 sweep of BCS70. Data was collected during a home visit conducted in 2016-18, which compromised of 50 minutes interviews (face-to-face, computer-assisted-personal-interview and computer-assisted-self-completion interviews) with a further 50 minutes of biomedical assessments performed by trained nurses.[20] Participants provided informed consent and the study received full ethical approval from NRES Committee South East Coast - Brighton & Sussex (Ref 15/LO/1446).

### Objectively measured Physical Activity and Sedentary Behaviour

Cohort members were asked to wear the activPAL thigh mounted accelerometer (activPAL3 micro; PAL Technologies Ltd., Glasgow, UK), to measure both sitting and physical activity levels. The device uses derived information about thigh inclination and acceleration to estimate body posture (sitting/lying and upright) and transition between these postures, stepping, and stepping speed (cadence).[21–25] ActivPAL was validated for measuring free-living sedentary behaviour against direct observation using an automated camera (Youden’s Index: 92.48 [87.26-97.70]). [26] The thigh mounted device helps overcome established limitations of a hip-worn accelerometer, which are known to misclassify sitting and standing. [18]

The activPAL was set to record at the sampling frequency of 20Hz in accordance with other wear-time protocols.[27] The device was positioned on the midline anterior aspect of the upper thigh by the research nurses during the home visit, and was waterproofed, in line with the manufacturer’s recommendations. During the seven-day consecutive wear protocol, the cohort members were instructed to wear the device when performing any normal activities of daily living including exercise, swimming and bathing, and instructed not to reattach the device if removed. Data were processed using freely available software that has been previously validated with an almost perfect agreement (*k*>*0*.*8*) and a median Kappa of 0.94 (https://github.com/UOL-COLS/ProcessingPAL/releases/tag/V1.0).[28] The software uses an algorithm to isolate valid waking wear data from sleep or prolonged non-wear, summarized elsewhere. [28] MVPA was calculated using a step cadence ≥100.[29] Valid data was classified as 10 hours of waking wear time for at least one day.

### Contextual Sedentary Behaviour

Cohort members reported on a variety of different contexts of SB. This included on average how many hours were spent viewing TV and internet usage on a typical week day. Furthermore, participants were asked to rate their level of occupational physical activity using four response options (sitting; standing, physically active; heavy manual work occupations).

### Depression

Cohort members were asked if they had seen a doctor regarding depression and if they had used anti-depressant medication. Data on the usage of prescribed antidepressants was obtained from medication dispensers during the nurse’s visit, using the British National Formulary edition 69 code. Depression diagnosis was indicated by a positive response to both enquiries given that anti-depressants may in some instances be prescribed for conditions other than depression. Study members were also asked about symptoms of depression using the Malaise Inventory.[30] This 9-item scale has acceptable psychometric properties when compared to clinical review of the interview ratings from an experienced psychiatrist, with a receiver operating characteristic score of 0.87 for both case level depression and any current affective disorder.[31] Overall, the Malaise Inventory has good validity within the adult general population with high levels of internal reliability over 10 years between ages 23 and 33 (0.77 and 0.80 respectively). It has also been shown to have a moderate stability score over a period of 10 years of *r=0*.*52*.[32] The Malaise Inventory was scored from 0-9, with a greater score denoting a greater severity of depressive mood. A Malaise Inventory Score of ≥4 was used to indicate the presence of elevated depressive symptoms. [33]

### Covariates

Covariates were identified *a priori* from existing literature and included sex (male or female), device waking hours wear time, cigarette smoking (never smoked, ex-smoker, occasional and daily smoker), alcohol consumption (never, monthly, weekly), body mass index (underweight [<18.5 kg/m^2^], normal weight [18.5-24.9 kg/m^2^], overweight [25.0-29.9 kg/m^2^], obese [≥30 kg/m^2^]), disability using European Union-statistics on income and living condition [EU-SILC] classification (none; some extent, severely hampered), education (no qualifications, secondary school education [GCSE and A-Levels and Diplomas], graduate education [BSc, MSc, PhD]). For analyses using device measures of sitting we additionally adjusted for device measured MVPA (categorised into roughly equal tertiles: ≤0.6, 0.6-0.9, ≥0.9 hours,), although in analyses using self-reported data we adjusted for self-reported frequency of weekly exercise bouts, consisting of 30 minutes or more (0-1, 2-3, 4-5,6-7 per week).

### Statistical Analyses

Characteristics of the sample population were described in relation to device measured sitting. Multiple Logistic Regression was used to estimate the association between SB and depression diagnosis or symptoms. Device-measured sitting was modelled using daily averages which were categorised into roughly equal tertiles (<8 hours; 8-10 hours; >10 hours). Self -reported SB was categorised into four roughly equal groups (0-<1 hours; 1-2 hours; 2-3 hours; >3 hours). Occupational activity was based on four groups (sitting occupation; standing occupation; physical work; heavy manual work). The models were adjusted for sex, device wear time, cigarette smoking, body mass index, alcohol consumption, disability, educational attainment and MVPA. All statistical analyses were performed in IBM SPSS Statistics 26.

## Results

A total of 6,562 (88% of those invited) cohort members consented to participate in the present study. After removal of 1114 participants with unusable activPAL data (nurse unable to initiate (n=102); lost in post (n=591); unable to download (n=421)), and missing covariates (n=742), the analytic sample for device measured sitting comprised 4,706 participants (52.4% Female). Participants declining to wear the device were more likely to be male, smokers, report poorer health, and be obese. [34] The analytical sample for self-reported SB comprised 6,727 participants (52.2% Female).

### Device-measured sample characteristics

Table 1 shows that participants in the highest sitting tertile were more likely to be male, obese, report a depression diagnosis and greater depressive symptoms, to be smokers, consume alcohol on a weekly basis, have a secondary education, to have more severe disability and record the lowest amount of device measured MVPA. The prevalence of depression diagnosis and symptoms were 6.7% and 17.0% respectively, (Pearson correlation *r=0*.*27* between depression diagnosis and symptoms).

**Table 1:**
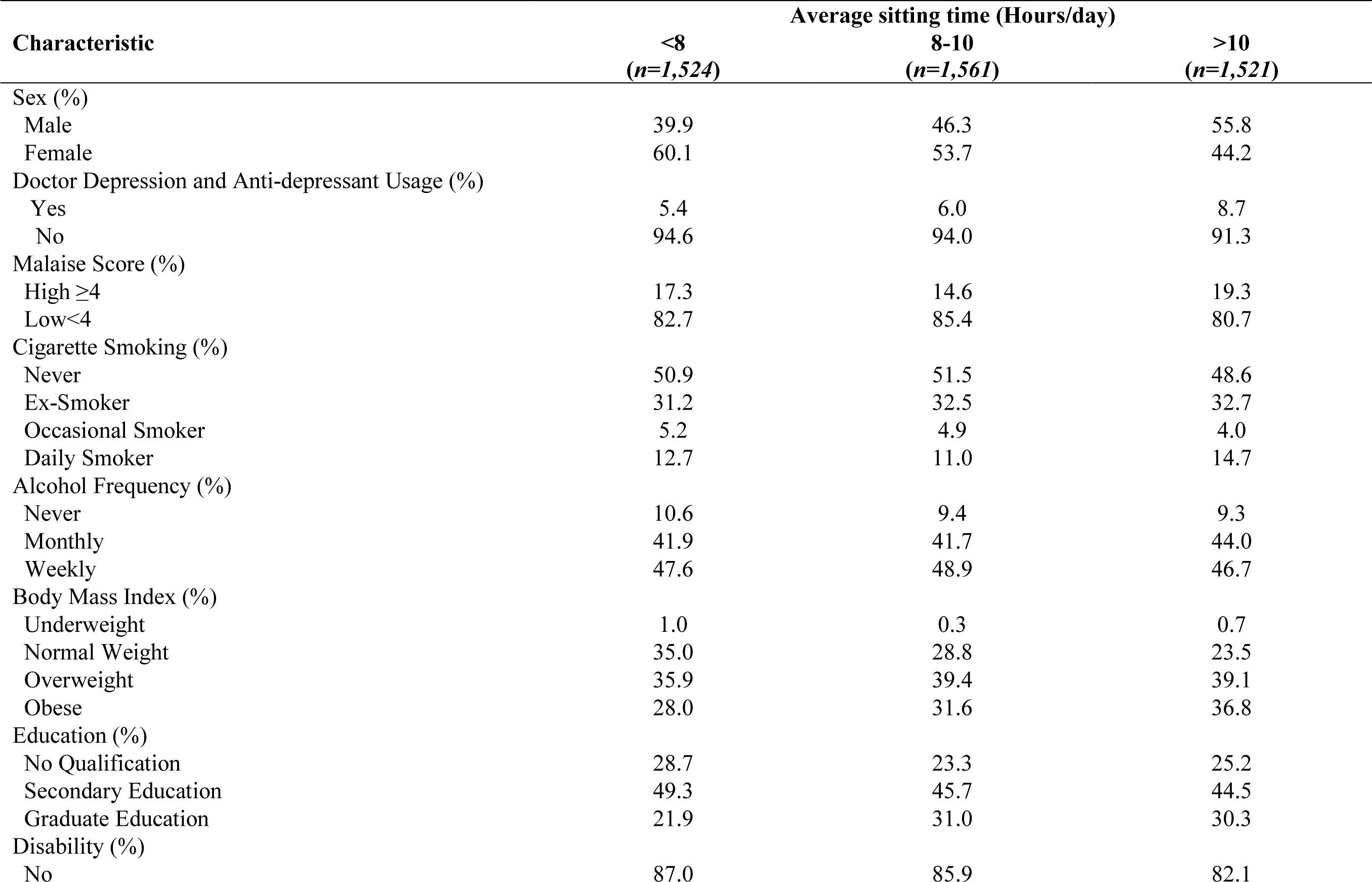
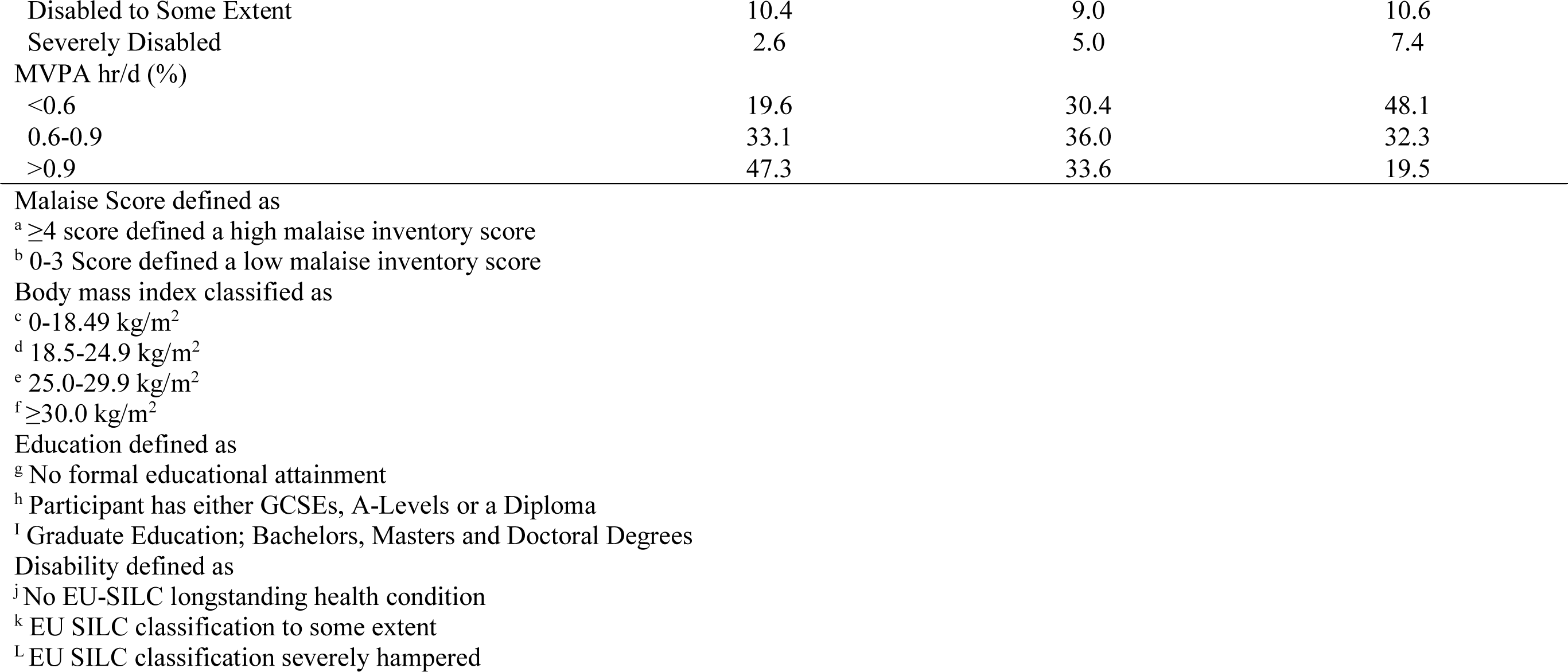
Characteristics of the study sample according to average sitting time at age 46, The British Birth Cohort Study

### Device measured sitting time, moderate-vigorous physical activity, sit to stand transitions and depression diagnosis

Compared to those who sat for <8 hours a day, participants who sat for longer hours had increased odds of depression diagnosis (Table 2). These associations persisted after adjustment for covariates albeit the effect estimate was attenuated. In contrast there were lower odds of depression diagnosis in participants recording >0.9 hours/day of MVPA, when compared to <0.6 hours/day (Table 2). However, no association was observed between device measured sit to stand transitions and depression diagnosis (Table S1).

**Table 2:** Device measured sitting time and moderate-vigorous physical activity in relation to depression diagnosis and symptoms, the British Birth Cohort Study

| **Doctor visit for depression and taking anti-depressants<sup>a</sup>** |  |  |  |  |  |
| --- | --- | --- | --- | --- | --- |
| **Total Sedentary Time**<br>**(Hours/day)** | **Yes**<br>**(n=316)** | **No**<br>**(n=4,388)** | **Model 1**<br>**OR (95%CI)** | **Model 2**<br>**OR (95%CI)** | **Model 3**<br>**OR (95%CI)** |
| <8 | 84 | 1,488 | 1.0 (Ref) | 1.0 (Ref) | 1.0 (Ref) |
| 8-10 | 97 | 1,479 | 1.31<br>(0.96-1.77) | 1.10<br>(0.80-1.52) | 1.04<br>(0.75-1.44) |
| ≥10 | 135 | 1,421 | 2.39<br>(1.77-3.23) | 1.67<br>(1.21-2.31) | 1.48<br>(1.05-2.08) |
| P-trend |  |  | <0.001 | <0.001 | 0.009 |
| **MVPA** |  |  |  |  |  |
| ≤0.6 | 172 | 1,362 | 1.0 (Ref) | 1.0 (Ref) | 1.0 (Ref) |
| 0.6-0.9 | 84 | 1,504 | 0.44<br>(0.34-0.58) | 0.73<br>(0.54-0.96) | 0.79<br>(0.58-1.07) |
| ≥0.9 | 60 | 1,522 | 0.32<br>(0.23-0.43) | 0.61<br>(0.44-0.86) | 0.70<br>(0.49-1.00) |
| P-trend |  |  | <0.001 | 0.001 | 0.017 |
| **Malaise inventory score** |  |  |  |  |  |
| **Total Sedentary Time**<br>**(Hours/day)** | **High≥4**<br>**(n=784)** | **Low<4**<br>**(n=3,815)** | **Model 1**<br>**OR (95%CI)** | **Model 2**<br>**OR (95%CI)** | **Model 3**<br>**OR (95%CI)** |
| <8 | 265 | 1,263 | 1.0 (Ref) | 1.0 (Ref) | 1.0 (Ref) |
| 8-10 | 226 | 1319 | 0.85<br>(0.70-1.03) | 0.80<br>(0.65-0.98) | 0.76<br>(0.62-0.94) |
| ≥10 | 293 | 1,233 | 1.27<br>(1.05-1.54) | 1.05<br>(0.85-1.30) | 0.95<br>(0.76-1.19) |
| P-trend |  |  | <0.001 | 0.305 | 0.955 |
| **MVPA** |  |  |  |  |  |
| ≤0.6 | 642 | 1,162 | 1.0 (Ref) | 1.0 (Ref) | 1.0 (Ref) |
| 0.6-0.9 | 241 | 1,313 | 0.61<br>(0.51-0.73) | 0.85<br>(0.69-1.04) | 0.85<br>(0.69-1.04) |
| ≥0.9 | 201 | 1,340 | 0.50<br>(0.41-0.61) | 0.75<br>(0.61-0.93) | 0.74<br>(0.59-0.93) |
| P-trend |  |  | <0.001 | 0.020 | 0.017 |

Model adjusted for
Model 1 – sex, device waking wear time (Hours)
Model 2 – sex, device waking wear time (Hours), cigarette smoking, body mass index, alcohol consumption, disability, education
Model 3 – sex, waking wear time (Hours), cigarette smoking, body mass index, alcohol consumption, disability, education, mutually for MVPA or sedentary
<sup>a</sup> Depression diagnoses was established via the combination of reported doctor visit for depression and reported taking a prescription of anti-depressants

### Device measured sitting time, moderate-vigorous physical activity, sit to stand transitions and depressive symptoms

There was some evidence of a ‘U’-shaped association between sitting and depressive symptoms, with participants reporting 8-10 hr/d, demonstrating lower odds of elevated symptoms (Table 2) compared to <8 hr/d of sitting time. MVPA was associated with lower odds of symptoms in a dose dependent manner (Table 2). Associations of sitting and MVPA were mutually independent across all statistical models. Conversely there appeared to be no association between total number of device measured sit to stand transitions and depressive symptoms (Table S1).

### Self-reported sedentary behaviours, depression diagnosis and depressive symptoms

The fully adjusted model showed no association between weekday TV viewing and depression diagnosis (Table 3). There was some evidence for elevated odds of depression with moderate (2-3hr/d) but not high (>3 hr/d) internet use (Table 4). No associations were seen for occupational activity and depression diagnosis (Table 5). Increased odds of depressive symptoms were observed in participants reporting >3 hours/day of TV weekday viewing time compared to <1 hr/d (Table 3). The association persisted in covariate adjusted models, albeit it was heavily attenuated. Both moderate and high internet use was associated with elevated depressive symptoms (table 4), although no associations were seen for occupational activity and depressive symptoms (table 5).

**Table 3:** TV weekday viewing time in relation to depression diagnosis and malaise inventory score, the British Birth Cohort Study

| **Doctor visit for depression and taking anti-depressants<sup>a</sup>** |  |  |  |  |  |
| --- | --- | --- | --- | --- | --- |
| **TV Weekday Viewing Time (Hours)** | **Yes (n=458)** | **No (n=6,269)** | **Model 1 OR (95%CI)** | **Model 2 OR (95%CI)** | **Model 3 OR (95%CI)** |
| 0-<1 | 58 | 1,017 | 1.0 (Ref) | 1.0 (Ref) | 1.0 (Ref) |
| 1-2 | 131 | 2,279 | 1.05<br>(0.76-1.44) | 0.99<br>(0.71-1.38) | 0.98<br>(0.70-1.37) |
| 2-3 | 123 | 1,743 | 1.26<br>(0.92-1.74) | 1.33<br>(0.80-1.60) | 1.13<br>(0.80-1.59) |
| >3 | 146 | 1,230 | 2.16<br>(1.57-2.96) | 1.31<br>(0.93-1.84) | 1.27<br>(0.90-1.79) |
| **Malaise inventory score** |  |  |  |  |  |
| **TV Weekday Viewing Time (Hours)** | **High (n=1,201)** | **Low (n=5,363)** | **Model 1 OR (95%CI)** | **Model 2 OR (95%CI)** | **Model 3 OR (95%CI)** |
| 0-<1 | 167 | 891 | 1.0 (Ref) | 1.0 (Ref) | 1.0 (Ref) |
| 1-2 | 385 | 1,977 | 1.06<br>(0.87-1.30) | 1.01<br>(0.82-1.25) | 1.00<br>(0.81-1.24) |
| 2-3 | 302 | 1,510 | 1.08<br>(0.88-1.33) | 0.97<br>(0.78-1.21) | 0.96<br>(0.77-1.20) |
| >3 | 347 | 985 | 1.92<br>(1.56-2.36) | 1.33<br>(1.22-1.61) | 1.28<br>(1.10-1.60) |

Model adjusted for
Model 1 – sex
Model 2 – sex, cigarette smoking, body mass index, alcohol consumption, disability, education
Model 3 - sex, cigarette smoking, body mass index, alcohol consumption, disability, education, self-reported exercise
<sup>a</sup> Depression diagnoses was established via the combination of reported doctor visit for depression and reported taking a prescription of anti-depressants

**Table 4:** Internet average weekday usage in relation to depression diagnosis and malaise inventory score, the British Birth Cohort Study

| Internet average<br>weekday usage<br>(Hours) | Doctor visit for depression and taking anti-depressants<sup>a</sup> |  | Model 1<br>OR (95%CI) | Model 2<br>OR (95%CI) | Model 3<br>OR (95%CI) |
| --- | --- | --- | --- | --- | --- |
|  | Yes<br>(n=456) | No<br>(n=6,267) |  |  |  |
| 0-<1 | 142 | 2,343 | 1.0 (Ref) | 1.0 (Ref) | 1.0 (Ref) |
| 1-2 | 168 | 2,468 | 1.14<br>(0.91-1.44) | 1.19<br>(0.94-1.52) | 1.19<br>(0.93-1.52) |
| 2-3 | 76 | 778 | 1.62<br>(1.21-2.17) | 1.52<br>(1.12-2.07) | 1.52<br>(1.12-2.07) |
| >3 | 70 | 678 | 1.72<br>(1.31-2.39) | 1.17<br>(0.84-1.62) | 1.14<br>(0.82-1.59) |
| Internet average<br>weekday usage<br>(Hours) | Malaise inventory score |  | Model 1<br>OR (95%CI) | Model 2<br>OR (95%CI) | Model 3<br>OR (95%CI) |
|  | High<br>(n=1,119) | Low<br>(n=5,363) |  |  |  |
| 0-<1 | 380 | 2,039 | 1.0 (Ref) | 1.0 (Ref) | 1.0 (Ref) |
| 1-2 | 427 | 2,152 | 1.08<br>(0.92-1.25) | 1.11<br>(0.95-1.30) | 1.11<br>(0.95-1.30) |
| 2-3 | 195 | 639 | 1.65<br>(1.35-2.00) | 1.60<br>(1.30-1.97) | 1.60<br>(1.30-1.97) |
| >3 | 197 | 533 | 2.04<br>(1.67-2.49) | 1.67<br>(1.35-2.07) | 1.63<br>(1.32-2.03) |

Model adjusted for
Model 1 – sex
Model 2 – sex, cigarette smoking, body mass index, alcohol consumption, disability, education
Model 3 - sex, cigarette smoking, body mass index, alcohol consumption, disability, education, self-reported exercise
<sup>a</sup> Depression diagnoses was established via the combination of reported doctor visit for depression and reported taking a prescription of anti-depressants

**Table 5:** Occupational work activity type in relation to depression diagnosis and malaise inventory score, the British Birth Cohort

| **Doctor visit for depression and taking anti-depressants<sup>a</sup>** |  |  |  |  |  |
| --- | --- | --- | --- | --- | --- |
| **Occupational work activity type** | **Yes**<br>**(n=300)** | **No**<br>**(n=5,390)** | **Model 1**<br>**OR (95%CI)** | **Model 2**<br>**OR (95%CI)** | **Model 3**<br>**OR (95%CI)** |
| Heavy Manual Work | 11 | 63 | 1.0 (Ref) | 1.0 (Ref) | 1.0 (Ref) |
| Sitting Occupation | 160 | 3,066 | 0.89<br>(0.47-1.69) | 1.09<br>(0.56-2.12) | 0.99<br>(0.50-1.94) |
| Standing Occupation | 40 | 872 | 0.79<br>(0.39-1.57) | 0.94<br>(0.46-1.12) | 0.86<br>(0.42-1.77) |
| Physical Work | 89 | 1,389 | 1.14<br>(0.59-2.18) | 1.32<br>(0.67-2.60) | 1.27<br>(0.65-2.50) |

| **Malaise inventory score** |  |  |  |  |  |
| --- | --- | --- | --- | --- | --- |
| **Occupational work activity type** | **High**<br>**(n=913)** | **Low**<br>**(n=4,940)** | **Model 1**<br>**OR (95%CI)** | **Model 2**<br>**OR (95%CI)** | **Model 3**<br>**OR (95%CI)** |
| Heavy Manual Work | 44 | 227 | 1.0 (Ref) | 1.0 (Ref) | 1.0 (Ref) |
| Sitting Occupation | 472 | 2,739 | 0.88<br>(0.47-1.67) | 1.08<br>(0.55-2.11) | 0.98<br>(0.50-1.92) |
| Standing Occupation | 141 | 768 | 0.78<br>(0.39-1.56) | 0.93<br>(0.45-1.92) | 0.85<br>(0.41-1.75) |
| Physical Work | 256 | 1,206 | 1.12<br>(0.58-2.14) | 1.30<br>(0.66-2.56) | 1.25<br>(0.63-2.45) |

Model adjusted for
Model 1 – sex
Model 2 – sex, cigarette smoking, body mass index, alcohol consumption, disability, education
Model 3 - sex, cigarette smoking, body mass index, alcohol consumption, disability, education, self-reported exercise
<sup>a</sup> Depression diagnoses was established via the combination of reported doctor visit for depression and reported taking a prescription of anti-depressants

## Discussion

The aim of the current study was to investigate the associations of device-measured sitting time and self-reported sedentary behaviours with depression utilising different methodological approaches. We found an increased odds of depression diagnosis in those with higher levels of device measured sitting time, although associations with context specific sedentary behaviours and depression diagnosis were less consistent. In contrast there was no association between device measured sitting time and depressive symptoms. TV viewing and internet usage were more consistently associated with higher depressive symptoms. This raises the possibility that the self-reported associations are spurious since depressive symptoms possibly bias the way in which participants respond to questions on sedentary behaviours. Device measured MVPA was associated with lower odds of both depression diagnosis and depressive symptoms in a dose dependent manner. There was no association between the number of device measured sit to stand transitions and depression (diagnosis and symptoms).

### Comparison to existing literature

Limited studies have utilised wearable devices to examine associations between sitting behaviour and depression in adults. Those that have were mostly conducted on small (n<500) convenience samples. Most of these studies [35-40], but not all [15] have noted associations between device measured sitting and higher risk of depressive symptoms assessed from self-reported questionnaires. To our knowledge only one small population based study [40] has previously employed the gold standard thigh worn position to examine sitting time in relation to depression. Our findings on device measured MVPA with both depression symptoms and diagnosis, supports the results from some previous studies [15,41], although others have only observed protective associations with light intensity activity and not MVPA [36-39]. It is conceivable that in older populations lighter intensity activity may be more beneficial forM mental health as greater exertion during vigorous forms of exercise may produce discomfort and shortness of breath, thus feel less enjoyable.

The data on TV viewing and internet usage are consistent with the results that were synthesised from a recent meta-analysis on the association between SB and diagnosed depression, which noted an increased risk of depression diagnosis in relation to TV viewing and computer usage.[11] Our finding of a null association between the number of sit to stand transitions and depression, appears largely consistent with a recent study on three elderly Scottish cohorts. Their data showed associations between sit to stand transitions and depression in only one of the three cohorts examined. [40] Indeed the mechanisms explaining any links between active breaks and depression seem unclear.

### Mechanisms

The possible explanation of an increased odds of depressive symptoms with both TV viewing and internet usage could be plausibly explained by factors such as social isolation, which is a potential cause for depression within the susceptible members of the population.[42,43] However, excessive prolonged periods of SB has also been associated with biomarkers of low grade inflammation, such as C-reactive protein. Thus, biological mechanisms may also explain associations between sitting and depression. This has been further supported by a randomised controlled trial, which observed higher depressive mood in parallel with elevated inflammatory biomarkers after a week of experimentally induced SB.[44]

### Strengths and limitations

The main limitation was the cross-sectional design as we cannot infer what direction the association is operating in. Bi-directional associations are not unlikely (i.e. whether sitting time is causing poor mental health or vice versa), although recent evidence using genetic instrumental variables suggested protective effects of physical activity on depression. [45] The depression outcome variables were self-reported, which could be influenced by reporting bias. Nevertheless, capturing anti-depressant usage has shown to be a valid approach for clinical depression diagnoses.[46]

A major strength of the current study is the combined use of devices and self-report tools to capture sedentary behaviour in its totality. This enabled us to capture both accurate and comprehensive information on total sitting volume and the context where such behaviours occur. The thigh worn position is considered the gold standard for free living sitting measurements. We adjusted our models for a wide range of covariates to limit possible residual confounding. Moreover, participants were all born in the same week of 1970, therefore limiting the confounding influences of age.

## Conclusion

Device measured sitting time was associated with clinically diagnosed depression. Self-reported sedentary behaviours were only associated with depressive symptoms. Encouraging physical activity and reducing sitting time could be beneficial in preventing depression.

## Data Availability

The datasets generated and/or analysed during the current study are publicly available from the UK data service https://www.ukdataservice.ac.uk/

## Declarations

### Ethics approval and consent to participate

Local research ethics committees approved all aspects of the survey (NRES Committee South East Coast - Brighton & Sussex, Ref 15/LO/1446) and all participants gave written informed consent.

### Consent for publication

Not applicable

### Availability of data and material

The datasets generated and/or analysed during the current study are publicly available from the UK data service <https://www.ukdataservice.ac.uk/>

### Authors’ contributions

MH/ES obtained funding, conceptualized and designed the study. AW performed analyses, drafted the initial manuscript. MH and GDB supervised the analyses and write-up. NP analysed Activpal data. All authors contributed to critical revision of paper and approved the final manuscript as submitted. MH is the manuscript’s guarantor.

## Acknowledgements

N/A

## Online Supplementary Tables

**Table S1:** Device-measured number of sit to stand transitions, in relation to depression diagnosis and malaise inventory score, the British Birth Cohort Study.

| Device-measured<br>number of sit to stand<br>transitions | Doctor visit for depression and taking anti-depressants <sup>a</sup> |  | Model 1<br>OR (95%CI) | Model 2<br>OR (95%CI) | Model 3<br>OR (95%CI) |
| --- | --- | --- | --- | --- | --- |
|  | Yes<br>(n=243) | No<br>(n=4,384) |  |  |  |
| 0-45 | 70 | 1,245 | 1.0 (Ref) | 1.0 (Ref) | 1.0 (Ref) |
| 46-60 | 88 | 1,705 | 0.91<br>(0.66-1.26) | 1.08<br>(0.77-1.53) | 1.08<br>(0.77-1.52) |
| 61-194 | 85 | 1,434 | 1.06<br>(0.76-1.49) | 1.38<br>(0.96-1.97) | 1.39<br>(0.97-1.99) |
| Device-measured<br>number of sit to stand<br>transitions | Malaise inventory score |  | Model 1<br>OR (95%CI) | Model 2<br>OR (95%CI) | Model 3<br>OR (95%CI) |
|  | High<br>(n= 678) | Low<br>(n= 3,915) |  |  |  |
| 0-45 | 200 | 1,110 | 1.0 (Ref) | 1.0 (Ref) | 1.0 (Ref) |
| 46-60 | 250 | 1,529 | 0.86<br>(0.71-1.06) | 0.94<br>(0.76-1.17) | 0.94<br>(0.76-1.16) |
| 61-194 | 228 | 1,276 | 0.93<br>(0.75-1.15) | 1.05<br>(0.84-1.31) | 1.05<br>(0.84-1.31) |

Models adjusted for
Model 1 – sex, device wear time (Hours)
Model 2 – sex, cigarette smoking, body mass index, alcohol consumption, disability, education
Model 3 - sex, cigarette smoking, body mass index, alcohol consumption, disability, education, activity performed at work
<sup>a</sup> Depression diagnoses was established via the combination of reported doctor visit for depression and reported taking a prescription of anti-depressants

